# Population-specific Risk of Pharmacogenomics-related Inaccurate Drug Dosing of ICU Patients

**DOI:** 10.1101/2025.02.11.25321889

**Authors:** Mahboubeh R. Rostami, Juan Rodriguez-Flores, Ali Ait Hssain, Alya Al Shakaki, Huda Khan, Muneera Vakayil, Edin Karic, Maha Elhamid, Lubna Gamal Al Tawil, Jason G. Mezey, Amal Robay, Ronald G. Crystal

**Affiliations:** Department of Genetic Medicine Weill Cornell Medicine, New York, New York; Department of Genetic Medicine Weill Cornell Medicine – Qatar; Medical Intensive Care Unit Hamad Medical Corporation, Doha, Qatar; Department of Biological Statistics and Computational Biology Cornell University, Ithaca, NY

**Keywords:** Intensive care unit, variants, whole genome sequencing, Qataris

## Abstract

**Rationale:** Intensive care units (ICU) patients are highly vulnerable to inaccurate drug dosing. Pharmacogenomics (PGx) characterizes the influence of inherited genetic variation on drug metabolism, playing an important role in the consequences of a given drug dose.

**Objectives:** To assess the genetic-based risk of inaccurate drug dosing in the ICU.

**Methods:** We carried out whole genome sequencing (WGS) of 210 Qataris in ICU care at Hamad Medical Corporation (HMC), Doha, Qatar and assessed the WGS for predicted deleterious variants of genes that metabolize 30 drugs commonly prescribed in the ICU.

**Measurements and Main Results:** Analysis of 210 Qatari ICU WGS identified 329 variants predicted deleterious associated with 85 genes known to affect metabolism of the 30 ICU drugs. Of the ICU patients that received the 5 most commonly prescribed drugs (warfarin, phenytoin, midazolam, vancomycin, levetiracetam), 93% had deleterious metabolism-related variants. Most (91%) patients carried at least one variant in a gene that that had the potential to affect the metabolism or activity of at least 1 drug that the patient received. Most patients had ≥14 deleterious variants of genes that affect the metabolism of administered drugs. Comparison of the deleterious variants related to metabolism of ICU drugs with African/African American and European populations revealed significant population specificity in ICU related PGx variants.

**Conclusions:** Together, these data suggest that population specific, pharmacogenomics based on the individual’s genome likely plays a significant role in effective, safe dosing in the ICU setting.

## Introduction

Accurate drug dosing of patients in the intensive care unit (ICU) presents a significant challenge. ICU patients are critically ill, with complex, acute multiorgan dysfunction superimposed on preexisting chronic diseases. ICU populations have a higher incidence of adverse drug reactions compared to other hospitalized patients (1–3). These patients are highly vulnerable to inaccurate pharmacologic dosing, resulting in insufficient, ineffective, or excessive dosing, resulting in drug toxicity (4, 5). Among the many reasons for inaccurate drug dosing in the ICU is pharmacogenomics, the influence of the patient’s genome on drug metabolism (6–10). Pharmacogenomics characterizes the effect of inherited genetic variation on drug metabolism, likely playing an important role in the consequences of a given dose of a drug regarding efficacy and toxicity (11–13).

With this background, the focus of this study is to assess the risk to pharmacogenomic-based inaccurate dosing of patients in the ICU. We hypothesize that the typical ICU patient is at a theoretical pharmacogenomic-based risk to inaccurate dosing, and this pharmacogenomic-based risk is population specific. To date, there has been no whole genome pharmacogenomic assessment of actual ICU patients, only limited studies of pharmacogenomics in the ICU based on analysis of a few genes, analysis of whole genomes of the general population of drugs used in the ICU and have been confined to analysis of only European populations (6–10).

To assess the risk of pharmacogenomic-based inaccurate drug dosing in an ICU population, we carried out whole genome sequencing in 210 patients admitted to the medical ICU of Hamad Medical Corporation (HMC) Hospital, the major in-patient facility in Doha, Qatar and analyzed the genomes for predicted deleterious variants of genes known to affect the metabolism of the drugs administered to these patients. For the analysis, we only assessed the genomes of patients with Qatari hereditary, thus eliminating population-related differences in the genomes of pharmacogenomic-related genes (14–21). The Qataris are a highly inbred population, and as has been observed in other inbred populations (22), there is high probability of identifying variants at an elevated allele frequency and homozygotes for a variants only found in a heterozygous state in most other populations. To assess whether the ICU drug-related pharmacogenomic predicted deleterious variants were representative of the Qatari population, we compared the frequency of the predicted deleterious variants in the ICU pharmacogenomic-related genes in the genomes of the 210 Qataris in the ICU to 14,669 whole genomes of the Qatari population (23–25). Finally, to evaluate population-specificity of the predicted deleterious variants in the ICU pharmacogenomic-related genes, we compared the predicted deleterious variants in the ICU pharmacogenomic-related genes in the Qataris to the predicted deleterious variants in the ICU pharmacogenomic-related genes in 34,029 European and 20,744 African/African Americans.

## Methods

### Study Population

HMC ICU patients (n=210) were recruited to participate in the study (Table I). The study population were all Qatari, as defined by the reported ethnicity of parents and grandparents. There were 53% males and 47% females, 59% <65 years old and 41% ≥65 years old. The average body mass index was 30.7 ± 8.9. The primary reason to be in the ICU are summarized in Supplemental Table I. The typical patient had moderate to severe ICU-related disease severity. The average time in the ICU was 19.7 ± 33.4 days.

**Table I.** Demographic Characteristics of the 210 ICU Patients in this Study.

| Characteristics | Number (%) | Mean±SD<sup>8</sup> |
| --- | --- | --- |
| Qatari<sup>1</sup> | 210 (100%) |  |
| Male | 111 (53%) |  |
| Female | 99 (47%) |  |
| Age |  |  |
| 18-64yr | 123 (59%) |  |
| 65+yr | 87 (41%) |  |
| BMI |  | 30.7 ± 8.9 |
| Time in ICU (days) |  | 19.7 ± 33.4 |
| Severity |  |  |
| SOFA Score<sup>2</sup> |  | 7.7 ± 3.7 |
| Glasgow Coma Scale<sup>3</sup> |  | 11.7 ± 4.4 |
| Total APS<sup>4</sup> |  | 14.4 ± 6.5 |
| Total AGE<sup>5</sup> |  | 3.1 ± 2.2 |
| Total CHP<sup>6</sup> |  | 3.5 ± 2.3 |
| APACHE<sup>7</sup> |  | 20.9 ± 7.8 |

<sup>1</sup> Parents and grandparents are Qatari.
<sup>2</sup> Sequential Organ Failure Assessment (SOFA) score is a scoring system that assesses the performance of several organ systems in the body (neurologic, blood, liver, kidney and blood pressure/hemodynamics) (41).
<sup>3</sup> Glasgow Coma Scale a neurological assessment tool that measures a person's level of consciousness after brain injury (42).
<sup>4</sup> Acute Physiology Score measures the physiological condition of critically ill patients in ICUs based on 12 clinical measures (43).
<sup>5</sup> Acute Gastroenteritis severity is a 20-point scale that measures the clinical signs of AGE, including the duration of diarrhea and vomiting, the maximum number of diarrhea stools and vomiting episodes in a 24-hour period, the maximum recorded fever, and healthcare provider visits and treatment. It is scored as mild (0–8), moderate (9–10), or severe (≥11) (44).
<sup>6</sup> Chronic Health Point Score a component of the Acute Physiology and Chronic Health Evaluation II (APACHE II) score, is a tool used to assess the severity of a patient's illness and prognosis in ICU (45).
<sup>7</sup> Acute Physiology and Chronic Health Evaluation Score quantifies based on a patient's history of severe organ system insufficiency or immunocompromised status.
<sup>8</sup> SD – standard deviation.

### Recruitment, Sample Collection and Sequencing

For each patient or patient surrogate who provided informed consent, DNA was extracted from a 200 μl aliquot of blood using the Qiagen DNeasy Blood and Tissue Kit (www.qiagen.com). DNA quality was verified by quantified using a QBit quantification system (Thermo Fisher) and verified to be of high molecular weight using an Agilent D1000 TapeStation system (www.agilent.com). Illumina Nextera sequencing library preparation (www.illumina.com) was conducted on the samples. Six samples were pooled and barcoded for sequencing in parallel on a single Illumina flowcell designed to produce 30x coverage depth for a single genome, with a targeted sequencing yield of 5x per individual genome. Sequencing was conducted on an Illumina HiSeq 4000 instrument to produce paired-end 2x150bp reads for each sample. Sequence data was converted to FASTQ format for genome analysis using bcl2fastq (www.illumina.com).

### Whole Genome Sequencing Identification of Variants

Genetic variation data for single nucleotide variants (SNVs) and short insertions and deletions (indels) were generated for each individual and the full cohort using an established analysis workflow shown to yield highly reproducible variant calls (26). Sequencing reads were mapped to the latest human reference genome assembly (GRCh38) with Burrows-Wheeler Aligner (27) followed by preparation of mapped reads for variant calling using the GATK pipeline (28), SNV and indel variant calls were made using HaplotypeCaller (29). Variant calls for each individual were then merged into a single VCF file for the entire cohort. Variant quality was assessed through depth of coverage analysis, using code developed by the research team (30).

Variants identified in the cohorts were annotated by RefSeq (31) database using SnpEff (32). Known and novel variants were classified by comparing them to the latest DbSNP release (currently build 151) (33). Potentially deleterious variants were predicted using CADD (34) and SIFT (35), and the list of variants was cross-checked in the ClinVar database to assess their relevance to disease (36).

Details regarding selection of ICU-related pharmacogenomic genes and variants, pharmacogenomic analysis, analysis of other Qatar, European and African/African American whole genome database and data availability can be found in Supplemental Methods.

## Results

### Genes Coding for Enzymes that Metabolize Drugs Used in the ICU

For the 30 drugs commonly prescribed in the ICU at HMC (Table II), using information in the PharmGkb (https://www.pharmgkb.org) database (Supplemental Tables II, III), we identified 171 genes known to modulate the metabolism of those drugs (Table II). For some of the drugs used in the ICU, many genes can influence the metabolism of the drug. For example, the metabolism of warfarin, can be influenced by 22 genes, fentanyl by 15 genes, morphine by 22 genes, and phenytoin by 15 genes. In contrast, nimodipine, angiotensin II, nitroprusside, dexmedetomidine and ranitidine are metabolized by only one gene product.

**Table II.**
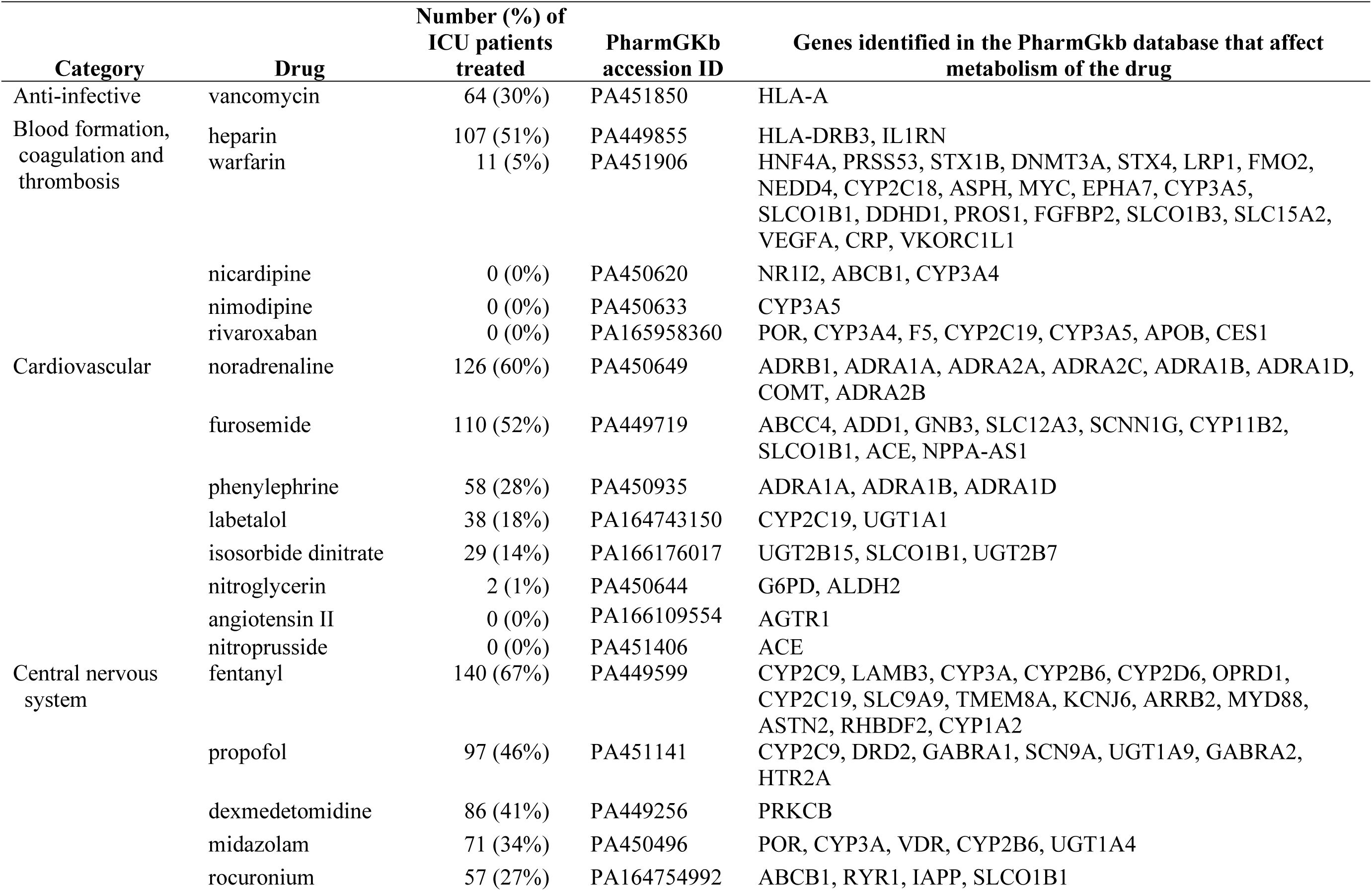
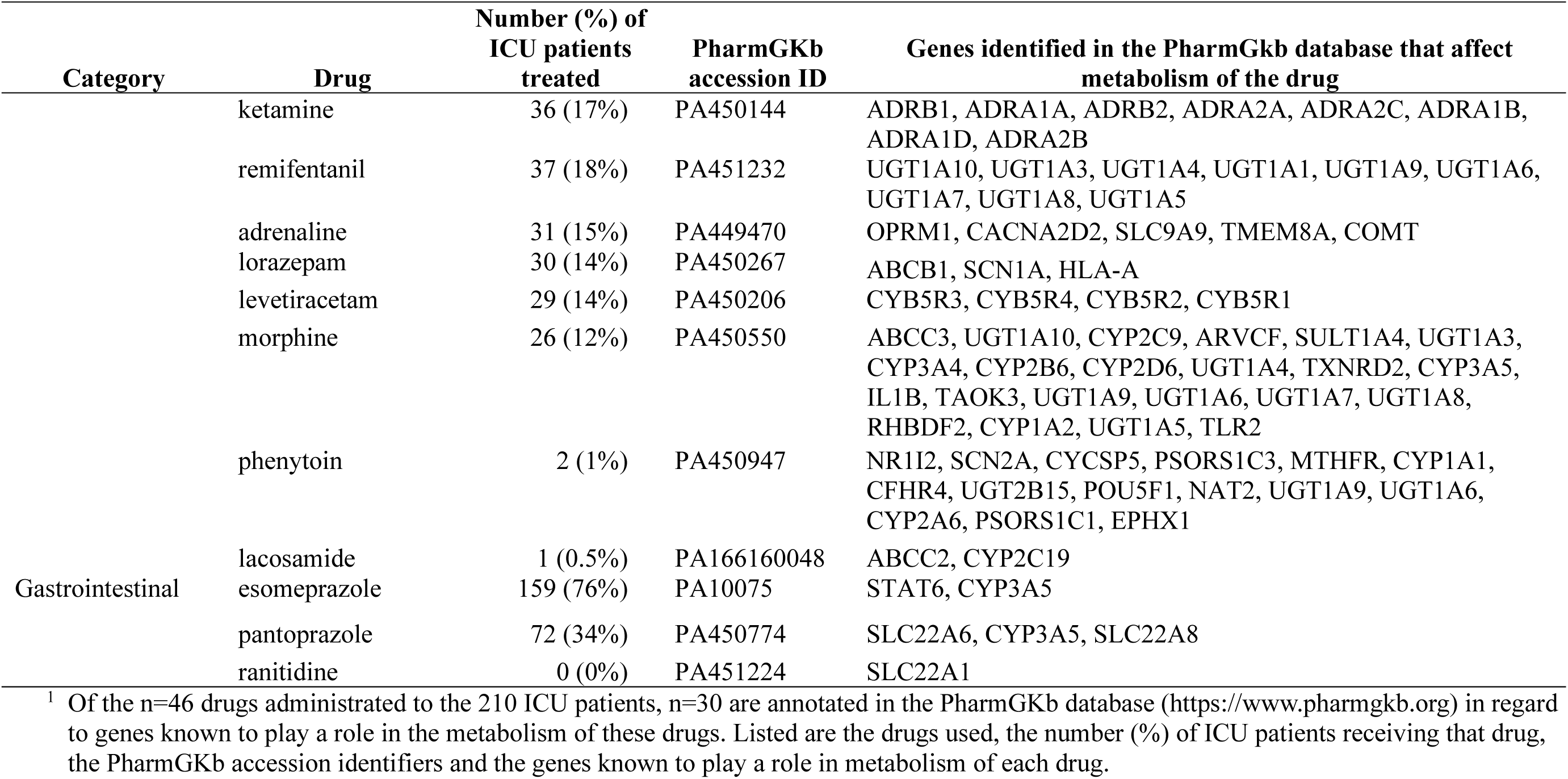
Drugs (n=30) Administered to 210 Patients in the ICU and the Genes (n=117) Known to Affect Metabolism of these Drugs Based on the PharmGKb Database^1^.

### Burden of Deleterious Variants of Genes That Metabolize Drugs Used in the ICU

For the 210 ICU patients, there was a total of 108,821 PGx variants of the genes relevant to metabolism of the 30 ICU drugs used in the ICU. The PGx variants identified as deleterious that would affect drug metabolism were identified based on SNPEff with high or moderate impact and CADD score ≥20 and SIFT <0.05. Of the 108,821 PGx variants, there were 329 deleterious variants associated with 85 genes (Supplemental Table IV). Of the 329 deleterious PGx variants, 46 (14%) were homozygous associated with 27 genes and 287 (87%) were known according to dbSNP (Table III). Based on allele frequency, 76 (23%) of the predicted deleterious variants were common, 37 (49%) were homozygotes and 39 (51%) heterozygotes.

**Table III.**
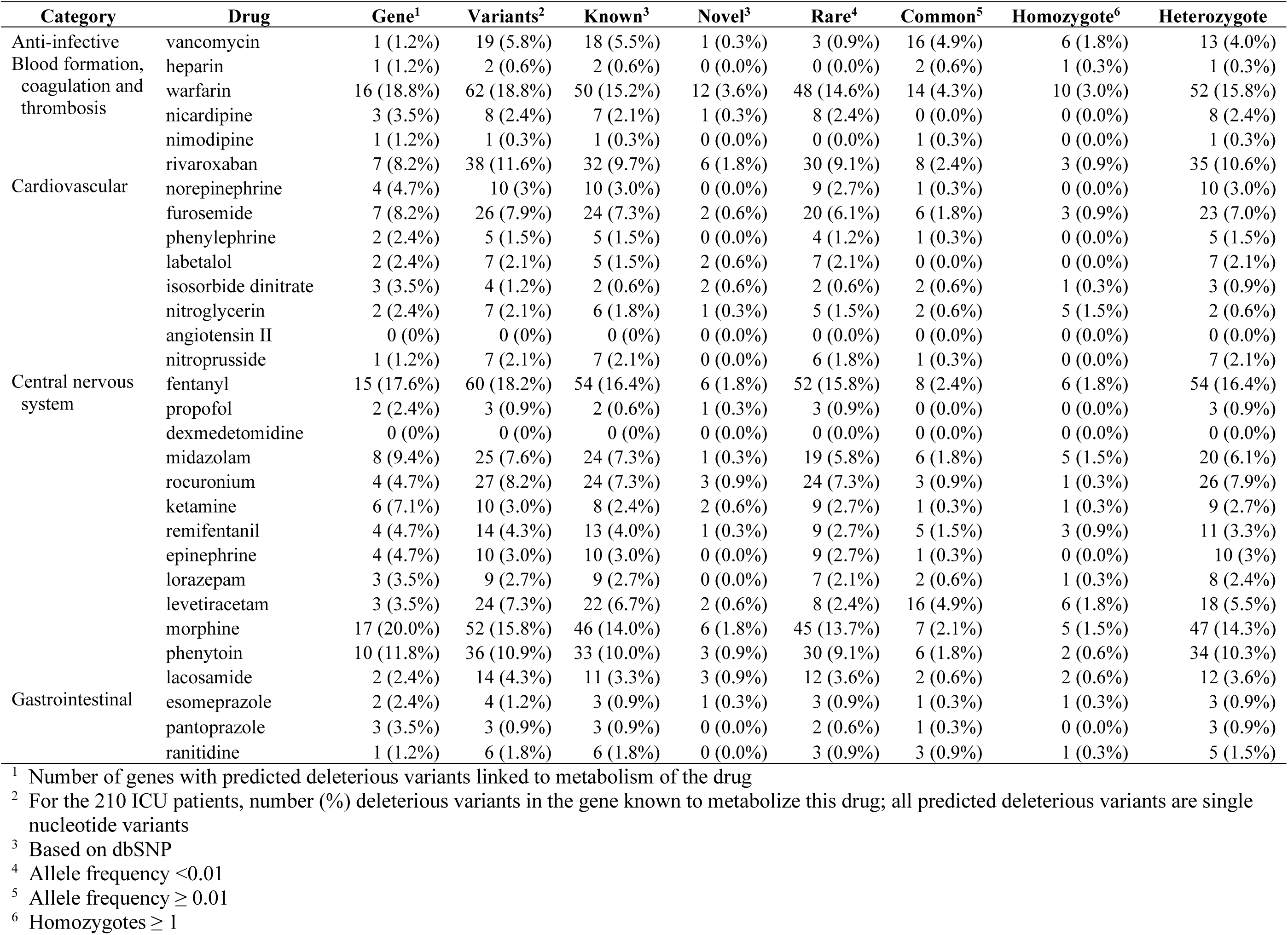
Number and Types of Deleterious Variants in Genes Known to Metabolize the Drugs Administrated to the 210 Qatari ICU Patients.

All ICU patients received at least 1 of 30 drugs commonly used drugs, and most received ≥5 of the 30 drugs (Figure 1A). The potential was high for an ICU patient having a deleterious variant of a gene effecting the metabolism of at least one of these drugs, with most patients having ≥9 genes with deleterious variants (Figure 1B). On a per patient basis, most ICU patients had a genomic burden of ≥14 deleterious variants of genes that affect the metabolism of administered drugs (Figure 1C) with patients receiving at least 1 drug for which that patient had a relevant deleterious variant (Figure 1D).

**Figure 1.**
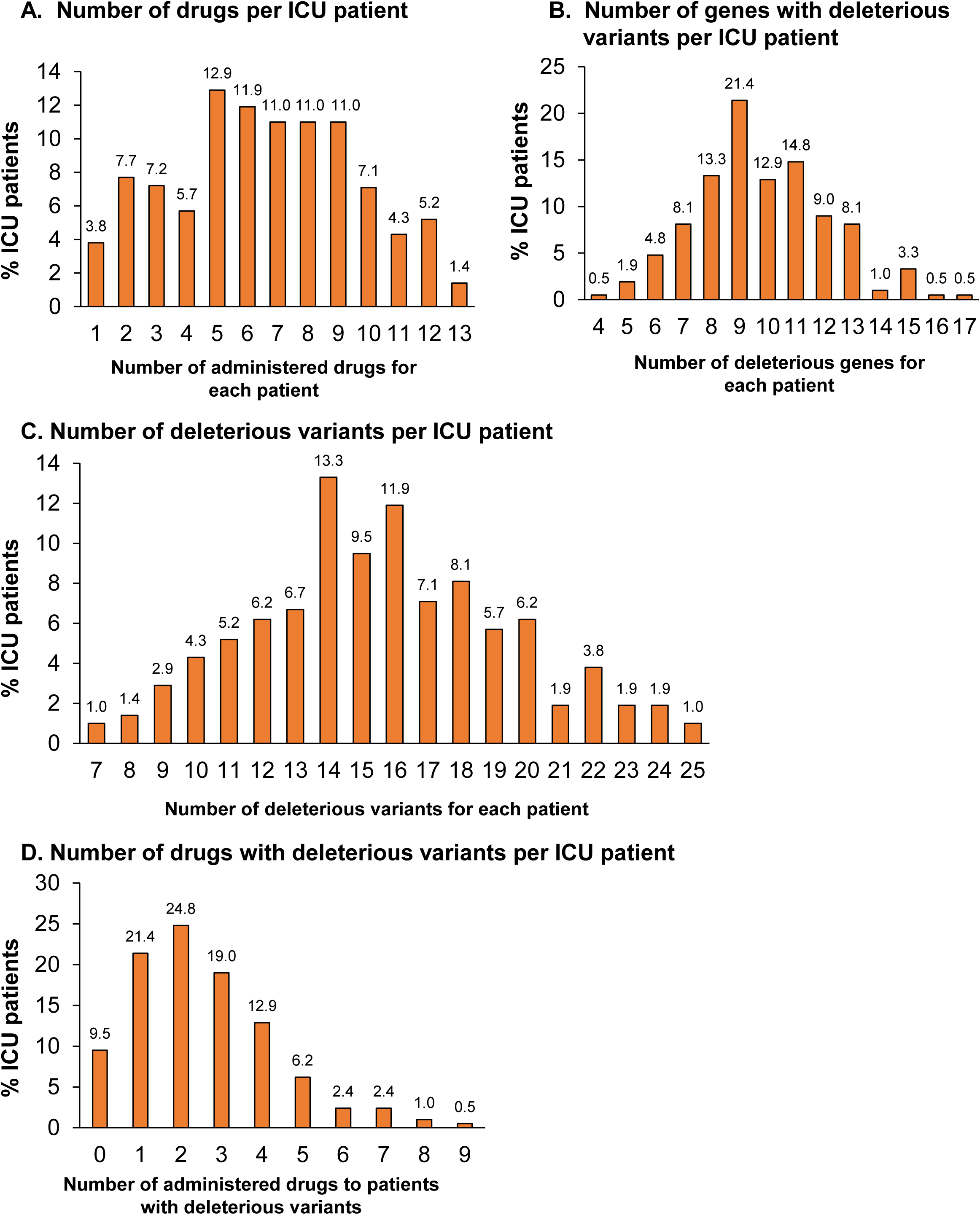
Burden per ICU patient of deleterious variants of genes known to play a role in the metabolism of drugs administered to the ICU patients. **A.** Average number of drugs administered to each ICU patient. The average ICU patient was administered 6.6 ± 3.0 different drugs while in the ICU. **B.** Average number of deleterious genes that metabolize the drugs administered to each patient; average 9.8 ± 2.3. **C.** Average number of deleterious variants in the drugs administered to each patient; 15.6 ± 3.8. **D.** Average number of administered drugs per ICU patient with deleterious variants for that drug; 2.5 ± 1.8. For all panels, data is presented as % ICU patients with the actual patient numbers above each bar.

### Drugs at Highest Risk for the Genome Influencing Metabolism of the Drug

Assessment of the number of predicted deleterious variants for each drug administered to each ICU patient identified levetiracetam, vancomycin, midazolam, fentanyl, rivaroxaban and warfarin at the highest risk for altered metabolism resulting from inherited deleterious variants of genes relevant to metabolism of these drugs (Figure 2A). The potential risk for altered metabolism of these drugs is high. For levetiracetam, vancomycin, warfarin, rivaroxaban, midazolam and fentanyl, each ICU patient had at least 1 predicted deleterious variant of one of the genes known to be relevant to the metabolism of that drug (Figure 1B). More than 93% of ICU patients carried at least one variant that could affect metabolism or activity of vancomycin or levetiracetam, and at least 50% of ICU patients carried at least one variant that could affect metabolism of furosemide, fentanyl, heparin, lorazepam, morphine and nitroglycerine (Figure 2C). For some drugs, including midazolam, heparin, warfarin, rivaroxaban, levetiracetam and vancomycin, the average allele frequency of deleterious variants was high (Figure 3). A significant number of ICU patients had >10 deleterious variants for drugs they received (Supplemental Figure 1).

**Figure 2.**
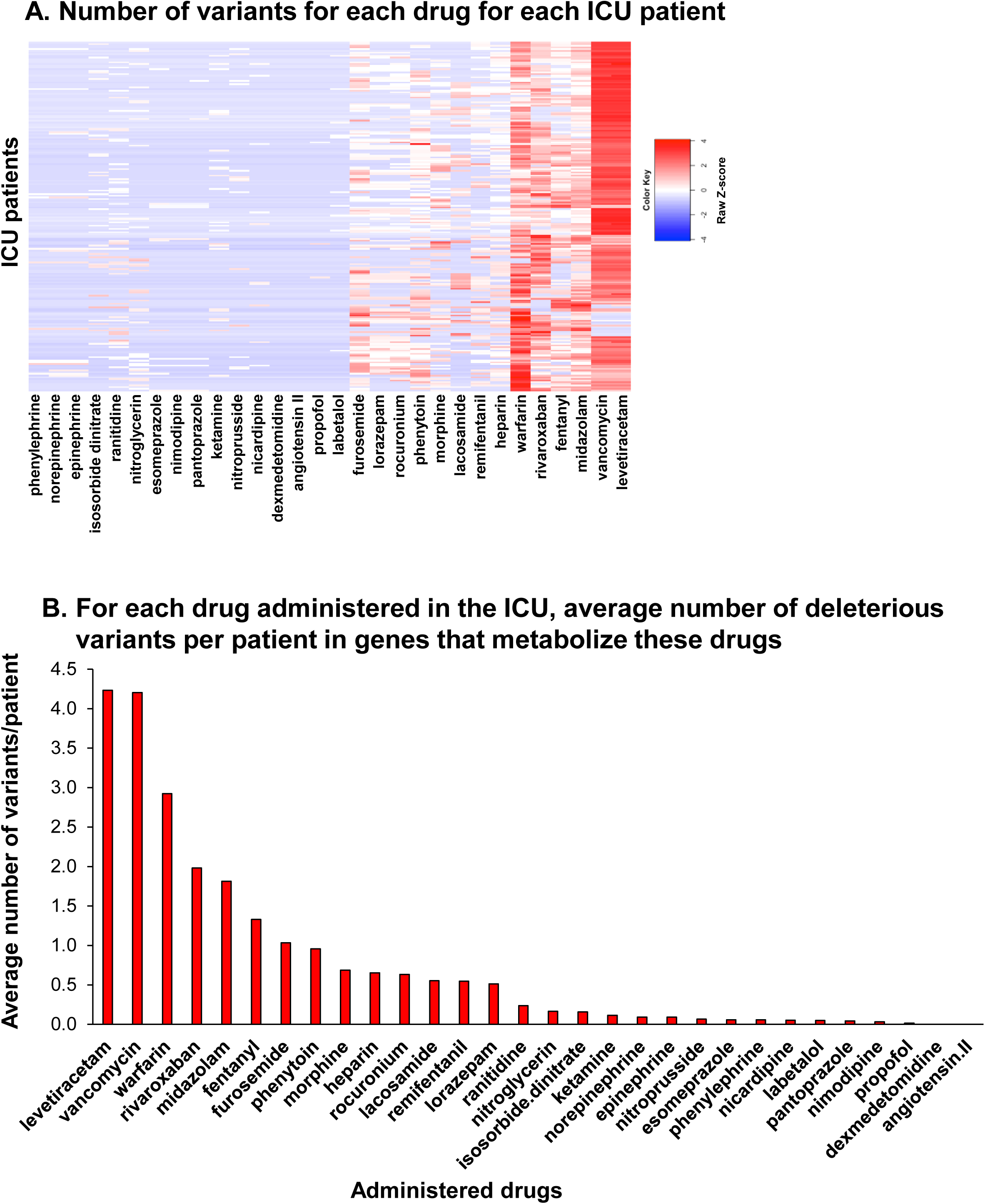
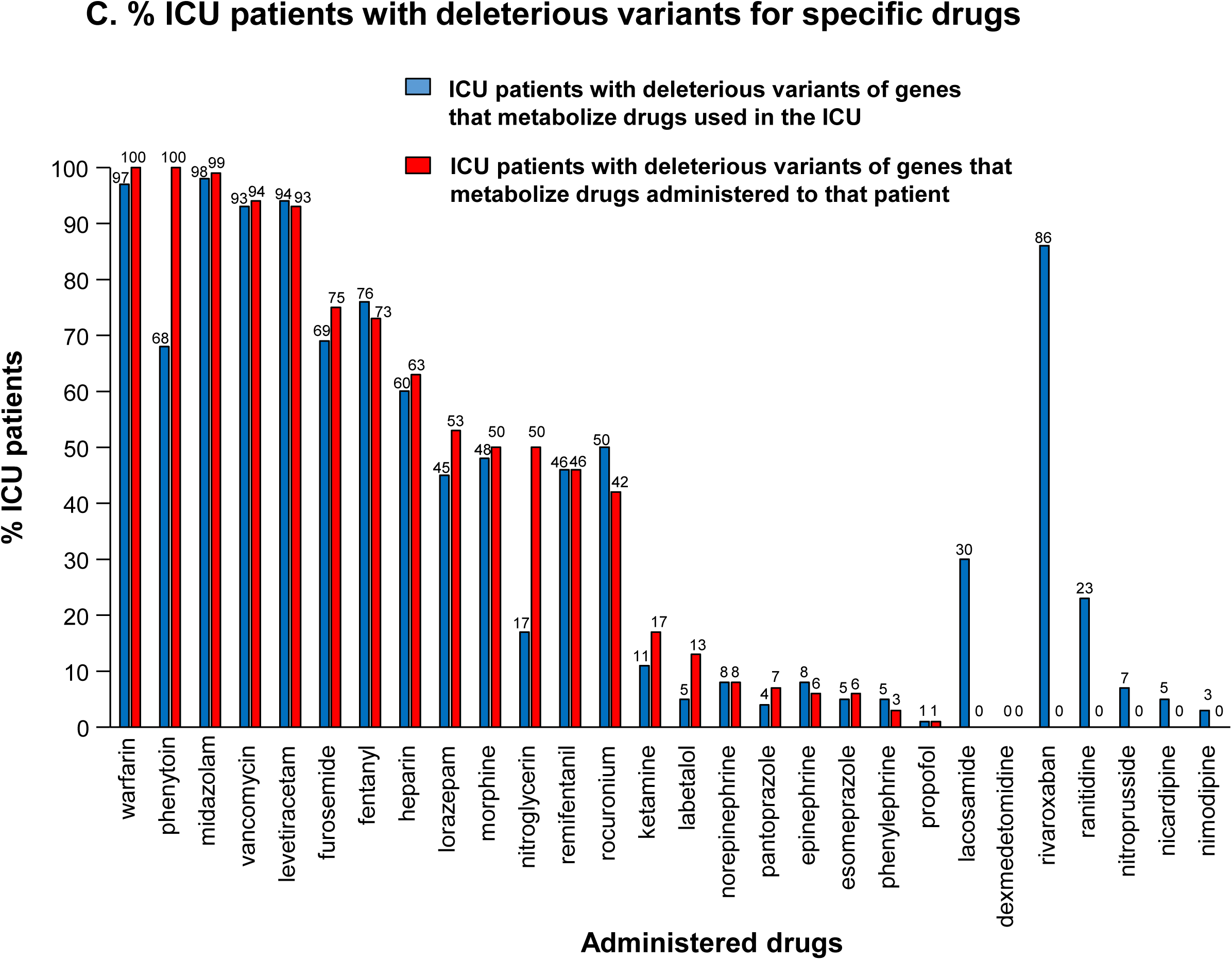
Burden for each ICU patient of deleterious variants of the drugs administered in the ICU. **A.** Heat map visualization of the burden of deleterious variants for each drug administered to each patient. Values of the heatmap are based on the Z-score (46). Blue lowest, red highest. Many of the ICU patients have deleterious variants in genes that metabolize levetiracetam, vancomycin, midazolam, fentanyl, rivaroxaban and warfarin. **B.** The average number of deleterious varians per patient in genes that metabolize each of the drugs administered in the ICU. For levetiracetam, vancomycin, warfarin, rivaroxaban, midazolam and fentanyl, on the average, each patient in the ICU had ≥1 deleterious variant in a gene that metabolizes these drugs. **C.** Percentage of ICU patients with deleterious variants in genes that metabolize specific drugs administered in the ICU (blue bars) compared to the actual % of ICU patients that received each drug.

**Figure 3.**
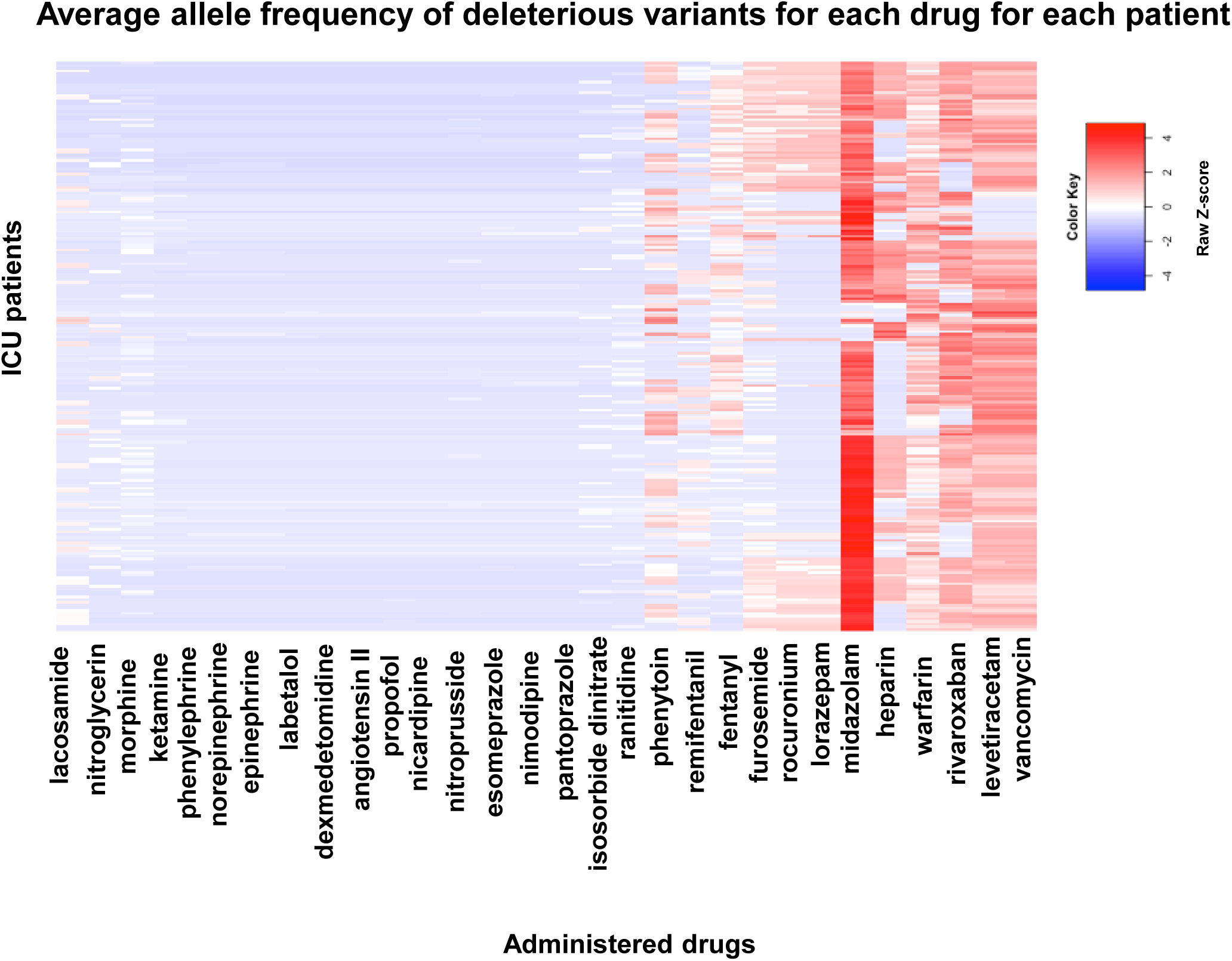
Heatmap of the average allele frequency of deleterious variants for each drug administered to each ICU patient. Based on the Z-score (46); blue lowest, red highest. The highest allele frequencies of deleterious variants was for vancomycin, levetiracetam, rivaroxaban, warfarin, heparin and midazolam.

### CYP Family Genes

By incorporating CYP Calling (37) into PharmCAT, we predicted the consequences of variants of the CYP family genes (Figure 4). For the CYP2D6 gene product, which metabolizes fentanyl and morphine, the predicted metabolizer categories among ICU patients included poor (1%), intermediate (19%), ultrarapid (4%), unknown (20%) and % normal (56%) metabolizers. Notably, 100 (48%) of the 210 Qatari ICU patients had deleterious variants that could impact morphine metabolism. Among the 26 (12%) ICU patients receiving morphine, 13 (50%) had deleterious variants affecting morphine metabolism.

**Figure 4.**
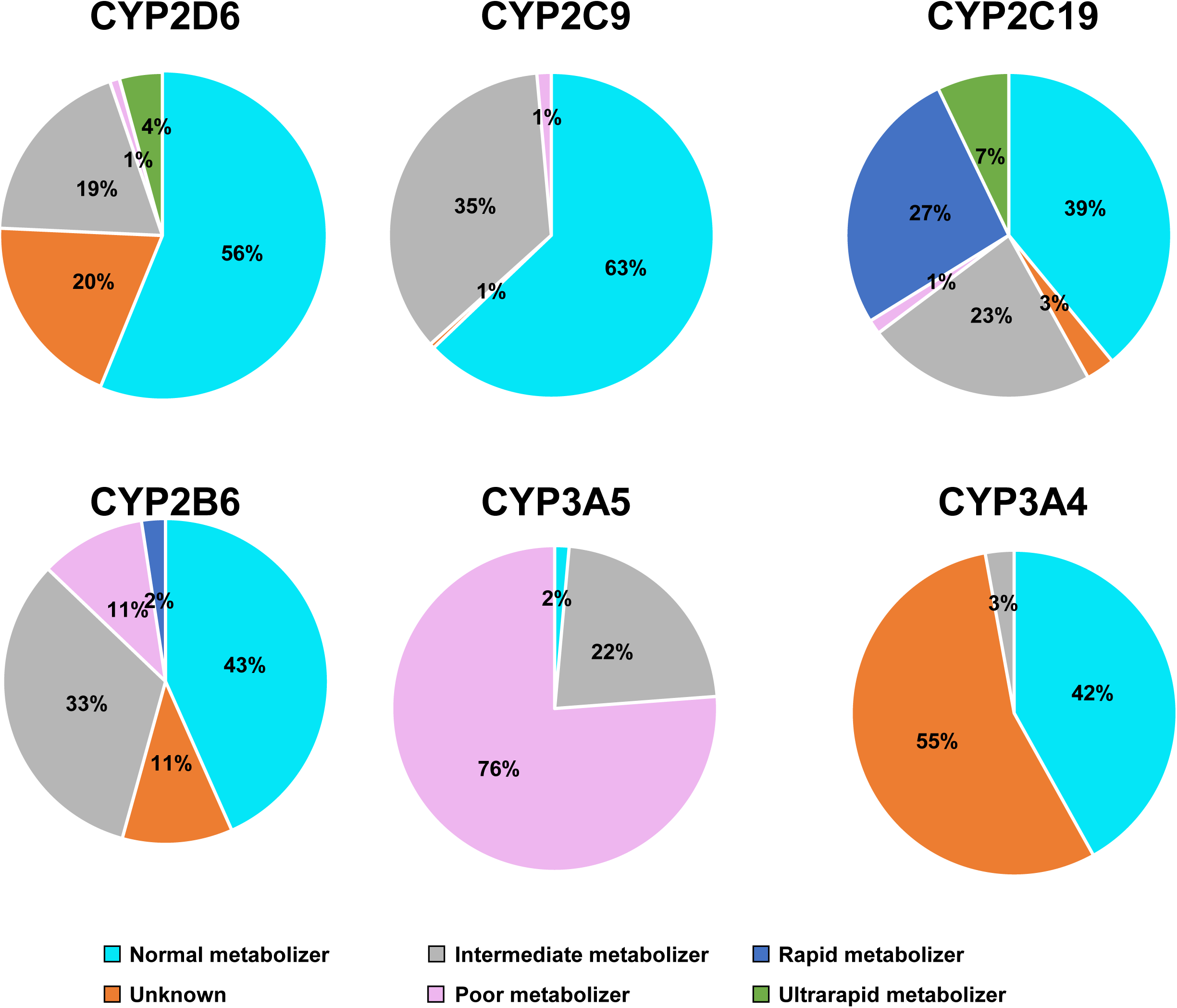
Examples of effects of deleterious variants of CYP genes on the extent of metabolism of drugs used in the ICU. The data is based on the whole genome sequences of the 210 in ICU patients and the annotations for each gene in the PharmGkb database. Shown for CYP2D6, CYP2C10, CUP3A5, CYP2C9, CYP2B6 and CYP3A4 are the allele frequencies for the extent of metabolism of drugs metabolized by the gene product of each CYP gene example. Light blue – normal metabolizer; grey – intermediate metabolizer; purple – rapid metabolizer; orange – unknown; pink – poor metabolizer; green – ultrarapid metabolizer.

For the CYP2C9 gene, involved in metabolizing fentanyl, morphine and propofol, 37% of ICU patients were classified as abnormal metabolizers. For the CYP2B6 genes which metabolize fentanyl, midazolam and morphine 57% of ICU patients were abnormal metabolizers. Regarding the CYP2C19 gene, which metabolizes labetalol, rivaroxaban, lacosamide and fentanyl, 61% of ICU patients were found to be abnormal metabolizers. For CYP3A4 which metabolizes rivaroxaban, morphine, nicardipine, fentanyl and midazolam, 58% ICU patients were abnormal metabolizer. For CYP3A5 which metabolizes fentanyl and midazolam 98% of ICU patients were abnormal metabolizers. Importantly, all ICU patients were abnormal metabolizers for at least for one gene from CYP family, with 52% of ICU patients were abnormal metabolizers for at least 4 CYP family genes (Figure 5, Supplemental Table VI).

**Figure 5.**
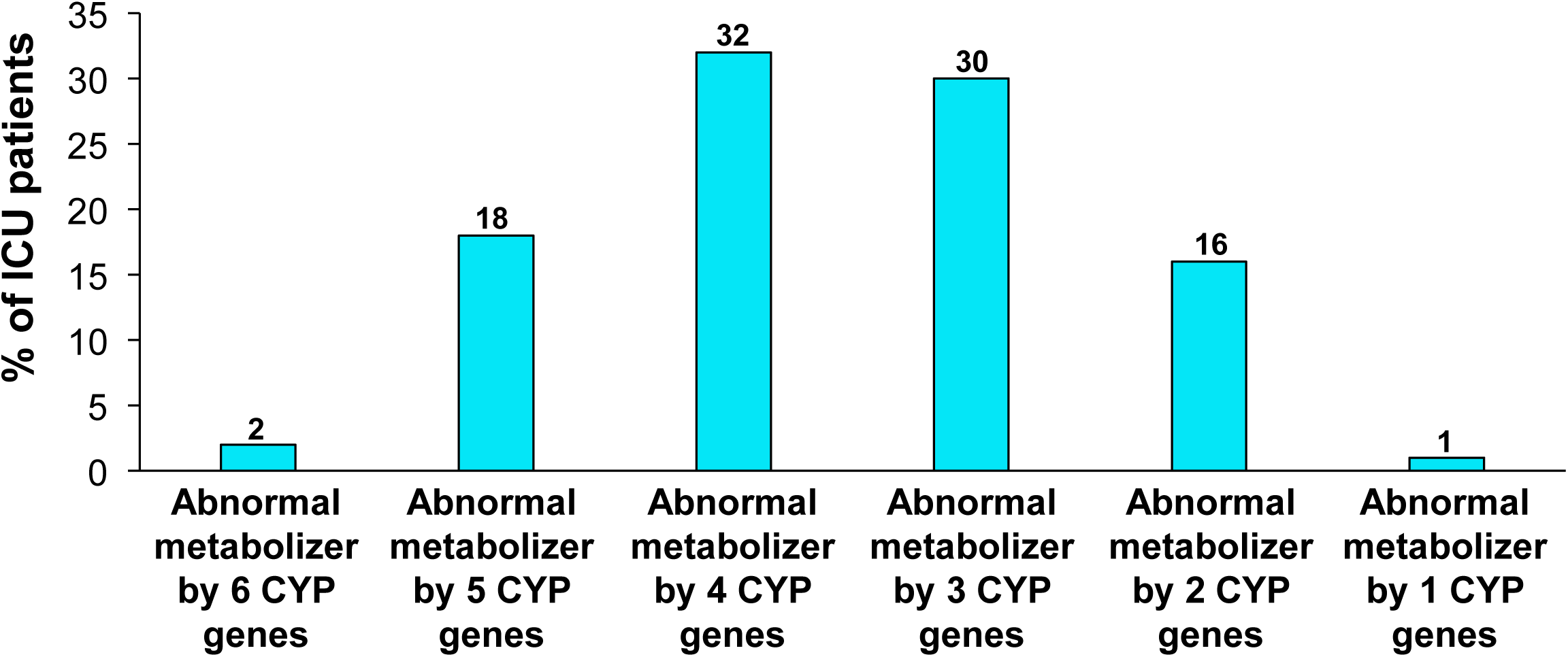
Burden if deleterious variants of CYP genes on metabolism of drugs administered to ICU patients. Based on the estimated deleterious variants in the whole genome of the 210 ICU patients and the PharmGkb database, the majority of ICU patients have variants of multiple CYP genes that metabolize drugs administered in the ICU.

### Comparison of PGx Variants in the ICU Patients with PGx Variants of the General Qatari Population

To quantify the impact of PGx screening on ICU care and assess the representation of these variants in the broader Qatari population, we compared the PGx variants of the 210 ICU patients related to the 30 commonly administered ICU drugs to 14,669 whole genome sequences (WGS) from the general Qatari population. In the QGP database, we identified 428,889 PGx variants of the 30 commonly used ICU drugs. Among these, 2,782 variants associated with 114 genes were classified as “deleterious.” Given the high rates of consanguinity within the Qatari population, we anticipated an increased occurrence of homozygous variants. Indeed, 252 variants (9%) were identified as homozygous in at least one of the 14,669 Qatari genomes analyzed. When comparing PGx ICU variants to those identified in the Qatari genome project (QGP) for administered drugs, we found significant overlap: 303 variants (92%) were shared between the two groups (Table IV). However, 26 variants (8%) were unique to ICU patients, highlighting the specific PGx considerations relevant to this setting.

**Table IV.**
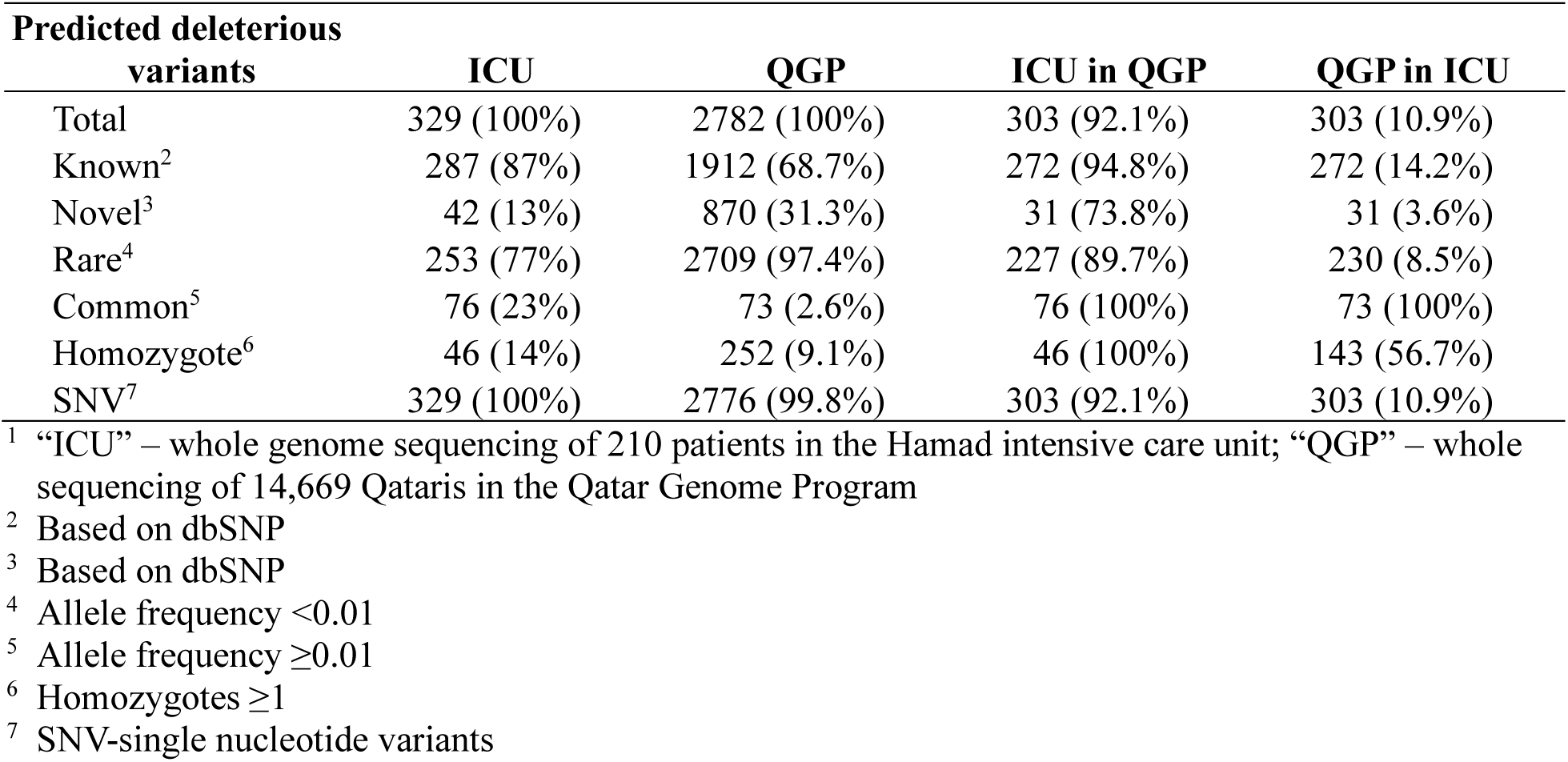
Comparison of the Genomes of the 210 ICU Patients to the Genomes of 14,559 QGP Qataris of Predicted Deleterious Variants in Genes that Metabolize Drugs Used in the ICU^1^.

### Comparison of PGx Variants Among Qatari, European and African/African American Populations

To assess the population specificity of the PGx-related genome variants of genes that metabolize the 30 commonly administered ICU drugs, we compared deleterious PGx variants among African/African American (n=20,744), European (n=34,029), and Qatari (n=14,669) populations. The total number of deleterious variants identified was 7,386 in the African/African American cohort and 11,980 in the European cohort. Of the Qataris, we identified 2,782 deleterious variants, of which 1,172 (42%) were also found in Europeans and 907 (33%) were present in Africans/African Americans. There was some overlap in PGx-related deleterious variants among the 3 populations, but there were striking differences. Overlap analysis revealed that unique variants comprised 1,531 (55%) of the Qataris, 7,203 (60%) of the European cohort, and 2,874 (39%) of the African/African American cohort (Table V). Notably, among the 907 variants identified in the African/African American population and Qataris, 261 (29%) exhibited allele frequencies that were 5 times higher than in the Qataris compared to African/African Americans, while in the European cohort, 645 (55%) of the 1,172 variants had 5X higher allele frequency in Qataris compared to Europeans.

**Table V.** Population Specificity of the Pharmacogenomic Deleterious Variants in Genes That Metabolize Drugs Administered in the ICU^1^.

| **Population<sup>2</sup>** | **Predicted deleterious variants** |  |  |  |  |
| --- | --- | --- | --- | --- | --- |
|  | **Total** | **Unique** | **Overlap in all 3 populations** | **Overlap with African population** | **Overlap with European population** |
| Qatari | 2,782 | 1,511 (55%) | 1,511 (55%) | 828 (30%) | 1,172 (42%) |
| African | 7,386 | 2,874 (39%) | 828 (11%) | – | 4,432 (60%) |
| European | 11,980 | 7,203 (60%) | 828 (7%) | 4,433(37%) | – |

<sup>1</sup> Comparison of predicted deleterious variants of the 30 drugs used to treat Qatari ICU patients at Hamad General Hospital to the predicted deleterious variants of the same genes in the European and African/African American populations.
<sup>2</sup> Datasets included: Qatari – QGP database (n=14,669); African/African American – gnomAD database (n=20,744); European - gnomAD database (n=34,029).

**Table VI.** Drug-specific, Population-specific Average Allele Frequency of Predicted Deleterious Variants of Genes that Metabolize Drugs Used in the ICU^1^.

| **Drug** | **Qatari** | **European** | **African/African American** |
| --- | --- | --- | --- |
| Midazolam | **0.22** | 0.05 | **0.22** |
| Fentanyl | 0.06 | 0.06 | **0.15** |
| Nitroglycerin | 0.05 | 0.00 | **0.12** |
| Isosorbide Dinitrate | 0.04 | 0.05 | **0.12** |
| Vancomycin | **0.11** | 0.05 | **0.11** |
| Levetiracetam | **0.11** | 0.05 | **0.11** |
| Warfarin | **0.10** | **0.10** | **0.10** |
| Heparin | **0.18** | **0.16** | 0.09 |
| Furosemide | 0.07 | **0.11** | 0.08 |
| Rivaroxaban | 0.11 | 0.02 | 0.07 |
| Remifentanyl | 0.07 | 0.07 | 0.07 |
| Ketamine | 0.02 | 0.03 | 0.05 |
| Morphine | 0.02 | 0.04 | 0.04 |
| Phenytoin | **0.14** | **0.10** | 0.03 |
| Ranitidine | 0.04 | 0.04 | 0.03 |
| Rocuronium | 0.07 | **0.15** | 0.03 |
| Lorazepam | 0.09 | **0.15** | 0.03 |
| Nicardipine | 0.00 | 0.03 | 0.02 |
| Labetalol | 0.00 | 0.00 | 0.01 |
| Lacosamide | 0.09 | 0.00 | 0.01 |
| Norepinephrine | 0.01 | 0.03 | 0.00 |
| Phenylephrine | 0.01 | 0.03 | 0.00 |
| Epinephrine | 0.01 | 0.03 | 0.00 |
| Nitroprusside | 0.02 | 0.00 | 0.00 |
| Esomeprazole | 0.00 | 0.00 | 0.00 |
| Propofol | 0.00 | 0.00 | 0.00 |
| Dexmedetomidine | 0.00 | 0.00 | 0.00 |
| Pantoprazole | 0.00 | 0.00 | 0.00 |
| Angiotensin II | 0.00 | 0.00 | 0.00 |
| Nimodipine | 0.00 | 0.00 | 0.00 |

<sup>1</sup> Drug-specific, population-specific comparison of the average allele frequency of the 30 drugs used to treat ICU patients in Hamad General Hospital; allele frequencies $\geq 0.10$ are bolded. The datasets include: Datasets included: Qatari – QGP database (n=14,669); African/African American – gnomAD database (n=20,744); European - gnomAD database (n=34,029).

### Differences in Allele Frequencies Among Different Populations

Not only were there differences in the predicted deleterious variants among the Qatari, African/African American and European populations, but there were significant drug-specific, population average allele frequencies of predicted deleterious variants of genes that metabolize midazolam was 22% for Qataris and African/African Americans but only 5% in Europeans. Significant population-specific allele frequencies were also noted for genes that metabolize vancomycin, levetiracetam, heparin, furosemide, phenytoin, rocuronium and lorazepam. This data highlights that for each drug, not only were there population differences in the deleterious variants of each gene, but there are significant allele frequencies among different ethnic/racial populations, i.e., each population has to be assessed independently.

## Discussion

ICU care requires administration of multiple drugs to a population that is highly vulnerable to inaccurate dosing, resulting in too little or to many drugs administered with consequent ineffective or toxic levels of the drug therapy (1–5). Assessment of predicted deleterious variants of drugs known to metabolize drugs commonly used in the ICU highlights the impact of genetic variations on drug metabolism of drugs commonly used in the ICU. Our study of whole-genome sequencing of a Qatari cohort in the ICU sheds light on the prevalence of deleterious variants in genes responsible for metabolizing drugs commonly used in the ICU setting. The important observation in this study is that variations in the genome of ICU patients have the potential to significantly alter the metabolism of drugs administered to ICU patients. This underscores the potential of pharmacogenomics to inform precision medicine approaches in intensive care, offering promising avenues for enhancing patient safety and treatment efficacy. Given the vulnerability of ICU to drug dosing (6–10, 38), identifying genomic variation of PGx variants of ICU patients could have significant impact on the quality of ICU care.

### Potential Impact of Genomic Variation on the Metabolism of Drugs Used in the ICU

By combining whole genome sequencing of ICU patients with the knowledge of the drugs administered to ICU patients which genes that metabolize these drugs it was possible to assess the potential influence of genomic variation on ICU dosing. The analysis led to several concepts. First, for many of the drugs commonly used in the ICU, there are many genes for which deleterious variants could significantly influence the metabolism of these drugs. Second, for many of these genes, there is a high allele frequency of deleterious variants of the PGx-related genes. Third, for each patient in the ICU, there is a high average of deleterious variants of PGx genes that effect metabolism of drugs administered to that patient. Fourth, on the average for each patient, there are several genes with metabolism-related deleterious variants. Fifth, of the 30 drugs commonly used in the ICU, there is a subset that are most likely affected. For Qataris, these drugs include warfarin, rivaroxaban, fentanyl, midazolam, vancomycin, levetiracetam, furosemide and heparin. Sixth, deleterious variants of the cytochrome P450 enzymes are common among the ICU patients and each patient commonly has inherited deleterious variants of several PGx-relevant CYP genes. Finally, the predicted deleterious variants of genes involved in metabolism of drugs used in the ICU are population specific in regard to genes, deleterious variants and allele frequencies, i.e., Qataris do not necessarily predict African/African Americans or Europeans and *vice versa*.

### Challenges to the Use of PGx Information in Guiding ICU Care

From the data from the present study, and prior PGx analysis of specific genes used in the ICU (7), a conclusion is that genomic analysis of PGx-related genes of ICU patients would be useful in guiding precision dosing in the ICU, with high metabolizers being administered higher doses and slow metabolizers lower doses. There are multiple challenges to putting this into practice.

First, there are significant differences in the genes/variants among different ethnic/racial groups as evidenced by demonstration of the significant differences among Qataris, African/African Americans and Europeans. Second, this is particularly critical in the ICU setting, where appropriate rapid therapy for critically ill patients is essential. Rapid availability of genotype diagnostic test results is crucial, along with clinicians’ ability to interpret these results for effective therapy. Third, pharmacogenomic-related parameters can be influenced by factors other than genomics, such as fluctuating blood volume, illness severity and both acute and chronic conditions affecting organ function. Most PGx studies have involved healthy volunteers or patients with single disease states, making it difficult to generalize findings to critically ill patients (7, 39). Fourth, the necessary polypharmacy in the ICU, where patients often receive multiple medications, complicates PGx interpretation due to potential drug-drug interactions (40).

In summary, whole genome sequencing of ICU patients compared to the PGx knowledge of the drugs commonly used in the ICU has demonstrated the high risk of the ICU patient to inaccurate dosing based on the individual’s genome of drug metabolizing genes. The challenge for the future is to develop a technology that can rapidly and accurately identify the genomic variants of the ICU patient to inform precision therapeutic dosing. For now, the data in the present study identifies specific drugs that are commonly used in the ICU and for which genomic analysis reveals a high probability that the encoding genes for enzymes that metabolize these drugs have variants that influence the metabolism of these drugs. Until the technology is developed to rapidly assess pharmacogenomic risk in the ICU, at a minimum, the identification in the present study of which ICU drugs are most likely affected by PGx variants can help guide the ICU staff to pay attention to the effectiveness and/or toxicity associated with these drugs.

## Acknowledgments

We thank all Qatar Research ICU Network collaborators : Abdelraouf Akkari, Awab El Shaikh, Ans Alamami, Abdulaziz Al-Alawi, Mohammed Qandil, Rabee Tawel, Ahmad Al Johari, Mohammed Faris, Anoud Duale, Khaled Ghazwi, Mojahid, Abdurrahman El Buzidi, Awadh Bin Taher, Ezzeddin Ibrahim and Nasseem Albadw for helping with patient recruitment, and N. Mohamed for editorial support. These studies were supported, in part, by Qatar Research Development and Innovation Council PPM 03-0314-190024 and Department of Genetic Medicine, Weill Cornell Medicine. The Qatar Biobank is funded by the Qatar Foundation for Education, Science and Community Development and the Supreme Council of Health.

## Supplemental Methods

### Selection of ICU-related Pharmacogenomic Genes and Variants

To identify genes known to play a role in the metabolism of drugs used in the ICU, a list of all drugs administered to the ICU patients was generated (Supplemental Table II). Of the 46 drugs administered to the 210 ICU patients, 30 are annotated in the PharmGkb database (www.pharmgb.org; Table II). These 30 drugs were used for all subsequent analyses. Relationships extracted from the PharmGKB annotations database (1) for the 30 ICU drugs totaled 587 associations between drugs and genes. After filtering out “ambiguous” or “not associated” (negative) associations, 587 remained, classified into 6 chemical/chemical, 171 chemical/gene, 139 chemical/haplotype and 271 chemical/variant associations (Supplemental Table III).

### PGx Analysis

The variants located in genes with a known drug metabolism role were extracted from the WGS of HMC ICU patients in Qatar. For a variant to be classified as “deleterious” it had to meet 4 criteria. First, the variant must have a high or moderate impact rate based on SnpEff annotation (2). Examples of “high impact” variants included frameshift, stop or start lost, transcript amplification, stop gained, splice donor or acceptor variants and transcript ablation. Moderate impact variants included in-frame insertion or deletion, missense, protein-altering and regulatory region ablation. Second, the variants were assessed for deleterious determined by “Combined Annotative Dependent Depletion” (CADD) with the criteria of “dysfunction” if the CADD score was ≥20 (3). Third, mis-sense variants had to meet the criteria of “Sorting Intolerant from Tolerant” (SIFT) analyses, with the variant not classified as “tolerant” (SIFT score ≥0.05). Fourth, the list of variants passing the first 3 criteria were cross-checked with ClinVar and any variant previously described as either “benign” or “likely benign” in ClinVar was removed. The predicted deleterious variants were then characterized as “rare” (allele frequency <0.01) or “common” (allele frequency ≥0.01; Supplemental Table IV).

### CYP Family Genes

PharmCAT (Pharmacogenomics Clinical Application Toolkit) ](https://pharmcat.org) was developed to extract PGx variants from sequencing or genotyping technologies, determine genotypes and diplotypes, infer corresponding phenotypes, and connect these with clinical prescribing recommendations from sources like the Clinical Pharmacogenetics Implementation Consortium (CPIC) and the PharmGKB-curated Dutch Pharmacogenetics Working Group (DPWG) (4). By incorporating CYP Calling (5) into PharmCAT, we predicted the consequences of variants of the CYP family genes.

### Qatar Genome Program (QGP), European and African / African American Whole Genome Databases

In addition to the whole genome sequencing of the 210 Qataris in the Hamad ICU, we compared the genes identified as related to metabolism of the 30 drugs in the ICU to the Qatar Genome Program (QGP) database of (n=14,669) whole genomes of Qataris. This enabled the answer to two questions, including: (1) how representative are the 210 ICU Qataris of the Qatari population in regard to PGx genes and variants; and (2) what are the population-specific differences in the deleterious variants relevant to metabolism of the drugs administered in the Hamad ICU? To answer the 1^st^ question, we compared to pharmacogenomic relevant variants in the 210 ICU participants to the pharmacogenomic-relevant variants in the QGP database. To answer the 2^nd^ question, we also assessed WGS of gnomAD (6)(https://gnomad.broadinstitute.org/ for v.4.1.0) publicly available database to identify deleterious PGx variants related to ICU medications in African/African Americans (n=20,744) and Europeans (n=34,029). We then compared the deleterious variants between these three groups (Supplemental Tables V-VII).

## Data Availability

The data supporting the finding of this study can be found in the main text or the supplemental information. Additional information may be made available upon request to the corresponding author.

## Supplemental Tables

Supplemental Table I. Reasons for Admittance to the ICU

Supplemental Table II. List of All Drugs Administered to the n=210 ICU Patients, With Identification of the n=30 Drugs Which Are Listed in the PharmGkb Database

Supplemental Table III. Relationships summarized (n= 587) from PharmGkb annotations database for 30 ICU drugs, Downloaded from PharmGkb

Supplemental Table IV. List of deleterious PGx variants (n=329) that metabolized 30 ICU drug in ICU (n=210)

Supplemental Table V. List of deleterious Pharmacogenomic variants (n=2,782) that metabolized 30 ICU drug in the QGP Database (n=14,669)

Supplemental Table VI. List of deleterious Pharmacogenomic variants (n=7,386) that metabolized 30 ICU drug in African/ African Americans (n=20,744)

Supplemental Table VII. List of deleterious Pharmacogenomic variants (n=11,980) that metabolized 30 ICU drug in European (n=34,029)

Supplemental Table VIII. CYP Deleterious Variants Consequences on Metabolism of Each ICU Patient

**Supplemental Figure 1.**
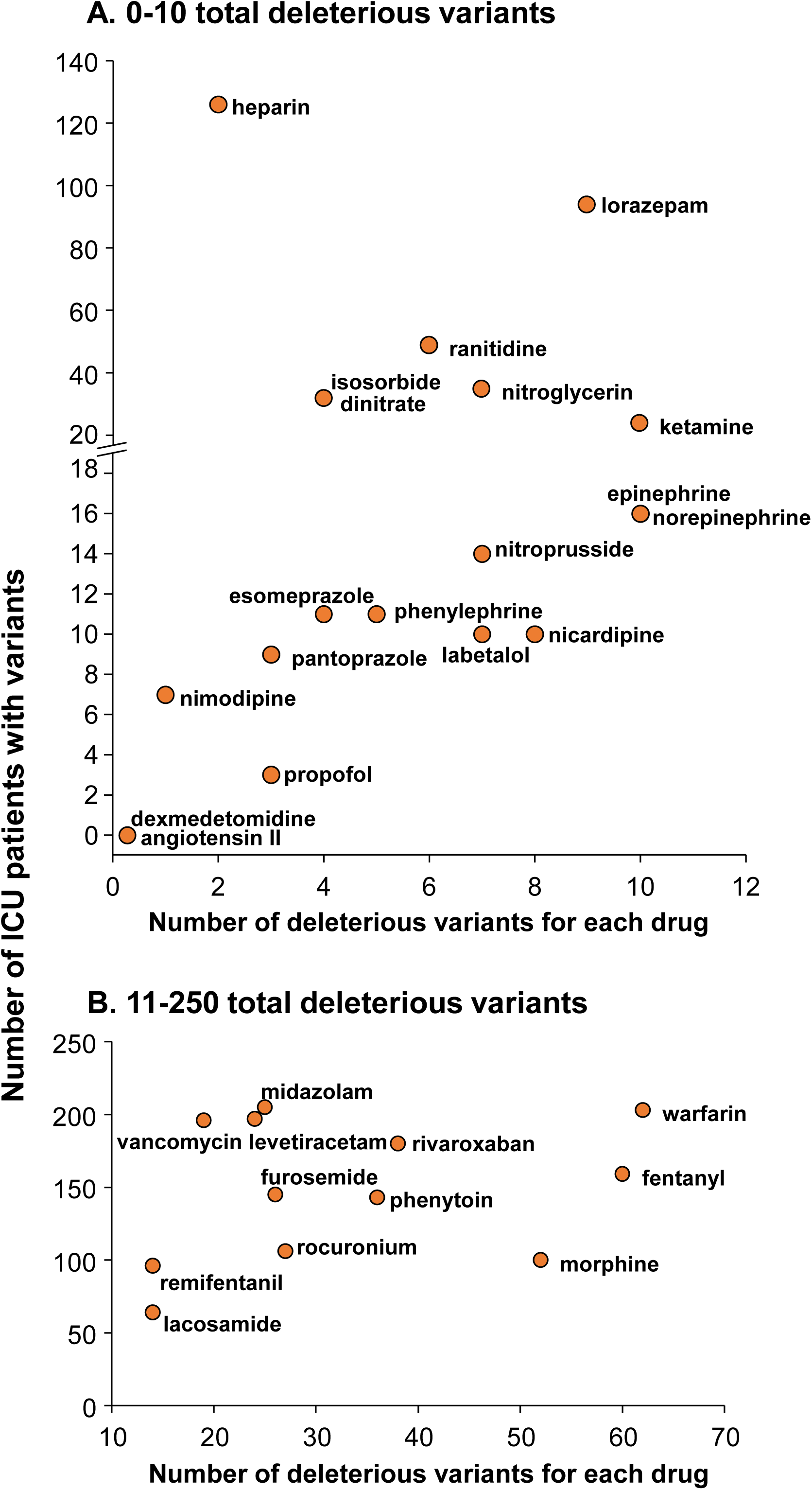
Number of ICU patients with deleterious variants for specific drugs administered in the ICU. *Top.* Number of ICU patients receiving drugs with 0-10 deleterious variants for each drug. *Bottom.* Number of ICU patients receiving drugs with 10-70 deleterious variants for each drug.

